# Evaluating General-Purpose LLMs for Patient-Facing Use: Dermatology-Centered Systematic Review and Meta-Analysis

**DOI:** 10.1101/2025.08.11.25333149

**Authors:** Irene S. Gabashvili

## Abstract

**Background:** General-purpose large language models (LLMs) have rapidly evolved from experimental tools into widely adopted components of healthcare. Their proliferation – accelerated by the “ChatGPT effect” – has sparked intense interest across patient-facing specialties. Among these, dermatology provides a high-visibility use case through which to assess LLM capabilities, evaluation practices, and adoption trends.

**Objective:** To systematically review and meta-analyze quantitative evaluations of general-purpose LLMs in dermatology, while extracting broader insights applicable to patient-centered use of AI across medical fields.

**Methods:** We conducted a multi-phase systematic review and meta-analysis, incorporating studies published through August 1, 2025. A total of 88 studies met inclusion criteria, covering over 100 dermatology-related tasks and yielding more than 2,500 normalized performance scores across metrics such as accuracy, sensitivity, readability, and clinical safety. This review also re-evaluates previously tested benchmarks to assess reproducibility and model improvement over time. Statistical analyses focused on heterogeneity (Cochran’s Q, I²), evaluator effects, and evolving methodological practices.

**Results:** LLM performance varied by architecture, prompt design, and task complexity. No single model demonstrated universal superiority, though retrieval-augmented and hybrid systems consistently outperformed others on complex reasoning tasks. Performance also varied by task, with smaller models sometimes outperforming flagships and “thinking” modes occasionally over-reasoning. Dermatology-specific models excelled in narrow contexts but lacked generalizability. Evaluation practices matured over time – shifting from static benchmarks to multi-rubric frameworks and simulations – yet high heterogeneity persisted (I² ≈ 90%) due to differences in study design and evaluator type.

Sentiment toward LLMs evolved from early skepticism (2022), to over-optimism (2023), to a more critical and diverse perspective by 2025. Preliminary ChatGPT-5 data, though limited to a small set of challenging conditions, suggest lower hallucination rates and better recognition of dermatological presentations on darker skin.

**Conclusions:** LLMs are entering clinical workflows rapidly, yet static evaluation methods often fail to keep pace. Our findings underscore the need for dynamic, modular, and evaluator-aware frameworks that reflect real-world complexity, patient interaction, and personalization. As traditional benchmarks lose relevance in the face of rapidly evolving model architectures, future evaluation strategies must embrace living reviews, human-in-the-loop simulations, and transparent meta-evaluation. Although dermatology serves as the focal domain, the challenges and recommendations articulated here are broadly applicable to all patient-facing fields in medicine.

**Limitations:** High heterogeneity, frequent model deprecation, and inconsistent study designs limit generalizability. While preliminary evidence from ChatGPT-5 shows improved performance for rare diseases and underrepresented skin tones, comprehensive, multi-model validation remains lacking. AI reliance on indexed literature continues to restrict the incorporation of patient-led research and independent evidence.

**Protocol Registration:** PROSPERO registration no. <u>CRD42023417336</u>

## Introduction

Over the past few years, general-purpose large language models (LLMs) have evolved from experimental tools to central components of our lives, increasingly integrated into healthcare research and clinical practice. This transformation, often described as the “ChatGPT effect”, has sparked widespread interest across medical specialties, especially in patient-facing fields like dermatology, which has historically embraced artificial intelligence (AI) innovations. However, as new models emerge and evolve at an unprecedented pace, traditional peer-reviewed evaluations frequently lag behind, diminishing their relevance by the time of publication. Dermatology, with its unique blend of visual diagnostics, consumer engagement, and openness to technology, offers a compelling case study for understanding how general-purpose LLMs are being assessed, adopted, and implemented in real-world clinical settings.

Since the first systematic review of LLMs in dermatology was published in 2023 [1], the scale and scope of relevant literature have expanded dramatically. This initial review laid the foundation for a living review, hosted on the Open Science Foundation (OSF) [2], designed to track the rapidly evolving landscape of general-purpose LLMs in dermatology. However, the pace of model development has far outstripped the evolution of robust evaluation methodologies – leaving the field with outdated benchmarks, inconsistent study designs, and limited guidance for assessing real-world utility. As of mid-2025, the living review has cataloged over 2,500 performance scores across 88 dermatology-focused studies [2], yet most of these assessments rely on static, one-turn prompt-response formats and evaluate models that have since been deprecated.

This updated review addresses these challenges by providing a longitudinal, meta-analytic synthesis of LLM evaluations in dermatology from 2022 to 2025. It not only captures trends in performance and sentiment but also critically examines the methodological frameworks that shape how LLMs are judged. A key emphasis is placed on model generalizability, clinical alignment, evaluator effects, and the shrinking “relevance window” of AI benchmarks. Through re-analysis of prior benchmarks, incorporation of model release timelines, and attention to nuanced shifts in sentiment and methodology, this review aims to set a new standard for evaluating general-purpose LLMs in dermatology and beyond.

## Methods

### Data Sources and Search Strategy

An extensive search was conducted across multiple databases, including EuropePMC, Semantic Scholar, PubMed, Dimensions AI, MedRxiv, BioRxiv, ArXiv, and Google Scholar, using keywords related to dermatology and AI (Appendix 1). Exclusion criteria were applied to filter irrelevant articles, and all matches were manually reviewed for relevance. For the live review, the number of papers retrieved from each source was recorded monthly to track and compare publication growth across platforms and fields.

### Protocol Registration

The study protocol was registered on PROSPERO (<u>CRD42023417336</u>). PRISMA guidelines were followed, with modifications to narrow the review’s scope.

### Study Selection

Studies were included based on relevance to dermatology and general-purpose language models. Exclusions included non-dermatological contexts. A team-based article summarization involved human and AI reviewers, with discrepancies resolved iteratively to minimize bias.

### Certainty and Risk of Bias Assessment

Evidence certainty was evaluated using the GRADE framework, accounting for risk of bias, inconsistency, indirectness, imprecision, and publication bias, supplemented by exploratory frameworks TRIPOD and CONSORT-AI. (Appendix 2)

### Date and Sentiment Labeling

Papers were labeled by both publication date and date of release of the most recent general purpose LLM used in the comparative analysis. This dual timestamping allowed us to distinguish between evaluations of current models and retrospective assessments of older ones. Papers that focused exclusively on non–general-purpose models (e.g., fine-tuned task-specific systems or LLM-based hybrids) were reviewed separately and excluded from the main sentiment and performance analysis. Since most studies reported multiple scores, each score was individually labeled by the release date of the exact LLM version responsible for that output. This fine-grained labeling enabled accurate aggregation of validity metrics and sentiment trends by model generation, rather than by publication lag.

Sentiment labels were derived through an iterative consensus-building approach that integrated outputs from multiple large language models (LLMs) with author-assigned sentiment labels. Initially, a set of published studies evaluating dermatological applications of LLMs was independently labeled by the author. Subsequently, three separate labeling sessions were conducted using different LLMs (GPT-4o, GPT-o4-mini, GPT-o3, Qwen3, Gemini 2.5). Each model was prompted to assign sentiment labels, either selecting from predefined categories (Positive, Satisfactory, Mixed, Cautionary, Contrasting, Negative, Unsatisfactory) or proposing novel classifications.

In cases of discrepancy or ambiguity – where models differed in their label assignments or suggested multiple plausible sentiments – a final adjudication step was performed. The author reviewed each model’s justification and reconciled disagreements by selecting the sentiment label that best reflected the consensus view across models and human assessment.

To test whether sentiment assignments differed systematically between LLMs and the human author, statistical comparisons (chi-square tests) were performed.

Detailed justifications and rationales supporting each final sentiment label decision, including explicit comparisons between human and model-assigned sentiments, are provided in Appendix 3.

### Data Extraction & Synthesis

Key metadata (e.g., DOI, publication type, MeSH terms, citation metrics) and study characteristics were extracted using Python scripts to support visualizations and synthesis of the dataset. Each paper was annotated with the following attributes: category, broader category, evaluation metrics and count, Fields of Research (ANZSRC 2020), Sustainable Development Goals, publication year, benchmark use, evaluation modality, presence of human expert or AI peer comparison, sentiment labels (final and alternate), reasoning complexity, knowledge depth, evaluation difficulty, and most recent model used with its release date.

For performance analysis, each reported metric was mapped to a composite variable termed Validity, normalized to a 0–1 or 0–100% scale depending on the original rubric. This encompassed accuracy, truthfulness, quality, and readability relative to expected standards. Additional fields included evaluation topic, dermatological condition, model version, sample size, and dataset description. Each score was tagged with the associated model version and either its release date or usage date, when available.

The dataset also includes re-evaluations of previously published benchmarks conducted as part of this review. These were used to assess reproducibility and detect performance drift over time.

For the meta-analysis, effect sizes were computed, and heterogeneity among studies was assessed using I^2^ and Cochran’s QQQ. A random effects model was applied to account for variability across studies. Additionally, subgroup analyses were conducted to explore patterns in specific study clusters, though the large number of potential subgroups necessitated careful prioritization. To perform fixed-effect meta-analysis across the most-frequently evaluated LLMs (each with ≥50 study entries), we converted “Validity (%)” into counts of “successes” (valid responses) based on each study’s sample size and computed the overall (pooled) validity proportion and its standard error under a fixed-effect model. For every LLM pair, we calculated the difference in pooled validity, its standard error, and a two-sided z-test. P-values were adjusted for multiple comparisons using the Benjamini–Hochberg procedure. One-way ANOVA and Kruskal-Wallis tests were performed to evaluate differences in accuracy and precision among LLMs. Pairwise statistical comparisons between model release years were conducted using Welch’s t-test, which accounts for unequal variances and sample sizes. Pairwise statistical comparisons of weighted mean validity scores between model release years using Welch’s t-test based on summary statistics (weighted mean, weighted standard deviation, and total sample size). Sensitivity analysis by excluding outliers was conducted to test the robustness of findings.

## Results

### Study Growth and Publication Trends

This review builds on our baseline [1], designed as a living systematic review with periodic updates on OSF [2]. The initial wave of literature followed shortly after the release of ChatGPT in late 2022, with hundreds of relevant studies published within six months. At that time, comprehensive domain-specific reviews, such as in dermatology, were still manageable in scope. In 2023, from 479 screened articles, 87 were selected (Figure 1). However, only one study featured a robust quantitative evaluation using six dermatology questions from Japan’s nursing board exam (then a preprint, now peer-reviewed), and three additional studies assessed responses to 1–2 questions (Appendix 2).

**Figure 1.**
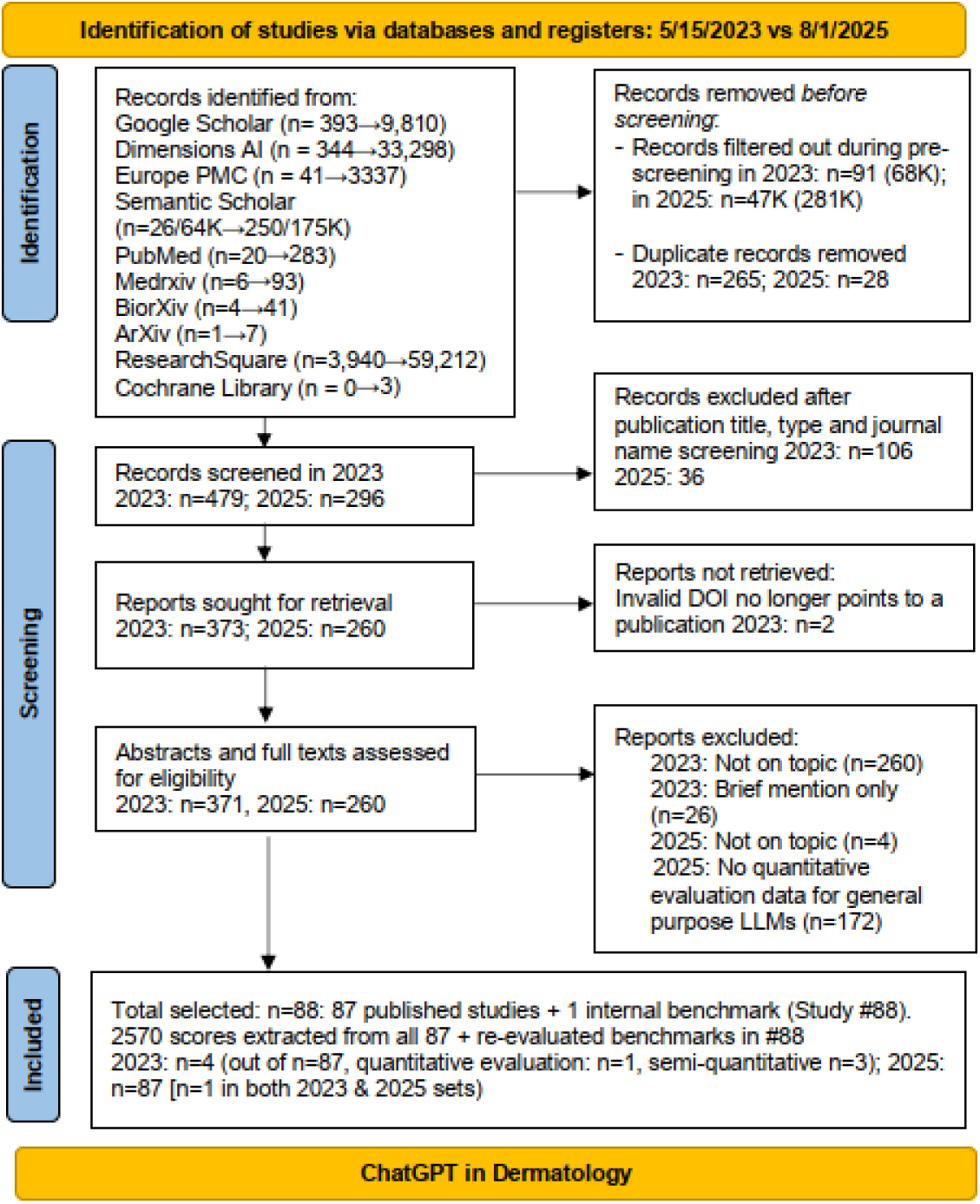
PRISMA Flow Diagram Comparing Study Identification (5/15/2023 vs. 8/1/2025)

By 2024, however, the scale of the literature required a refined strategy. We narrowed our scope to general-purpose LLMs with quantitative dermatology components, relying on titles, abstracts, and focused keyword filters. This yielded 1,700+ extracted scores from 56 studies. The upward trend continued in 2025, with 88 papers contributing more than 2,500 scores (Figure 1). We designate this study as #88, as it includes not only a systematic review but also re-evaluations of established benchmarks (see Appendix 3). Publication trends varied substantially by platform. Dimensions AI and Semantic Scholar demonstrated more faster growth than PubMed and MedRxiv, whose expansion was steadier but more modest.

Average monthly growth rates were ∼8.7% for PubMed and ∼10.2% for medRxiv, with both series showing substantially slower point estimates than larger non-indexed aggregators. However, when modeled jointly with other datasets (including Dimensions.AI, Europe PMC, and Google Scholar), differences in growth rates were not statistically significant in a global test (Wald χ² = 5.88, p = 0.44) or in pairwise comparisons (all p > 0.27). This suggests that, in relative terms, dermatology-specific literature is not clearly lagging behind other PubMed-indexed or preprint trends over the observed period.

A notable shift in publication venues occurred as general-purpose LLMs gained popularity. In 2023, the majority of related papers were released as preprints, with fewer than 10% appearing in specialized journals. By 2024, as the field matured, this trend reversed: the proportion of preprints declined to 7-12%, while a growing number of evaluations were published in peer-reviewed dermatology and biomedical journals.

Interestingly, early papers often acknowledged AI assistance (or named ChatGPT as co-author), but this practice declined in 2024, possibly reflecting increased caution or stigma around AI reliance.

Despite this surge in publications, 74% of scores in our dataset were from already discontinued models such as ChatGPT 3.5-4.0, Claude 1 and 2, Bard, LLAMA2, Gemini 1–1.5, and ChatGPT-o1. With the launch of ChatGPT 5 on August 7 2025, that share rose to 87%. This underscores how the pace of innovation has compressed the “relevance window” of state-of-the-art models, posing challenges for longitudinal tracking and reproducibility.

### Sentiment and Performance Over **Time**

Figure 2 illustrates the evolution of sentiment in LLM evaluation studies, grouped by the release year of the newest model evaluated in each paper. Sentiment was coded at the study level, reflecting the overall interpretation and tone rather than individual benchmark outcomes. Categories include *Positive*, *Satisfactory*, *Negative*, *Cautionary*, and *Contrasting*.

**Figure 2.**
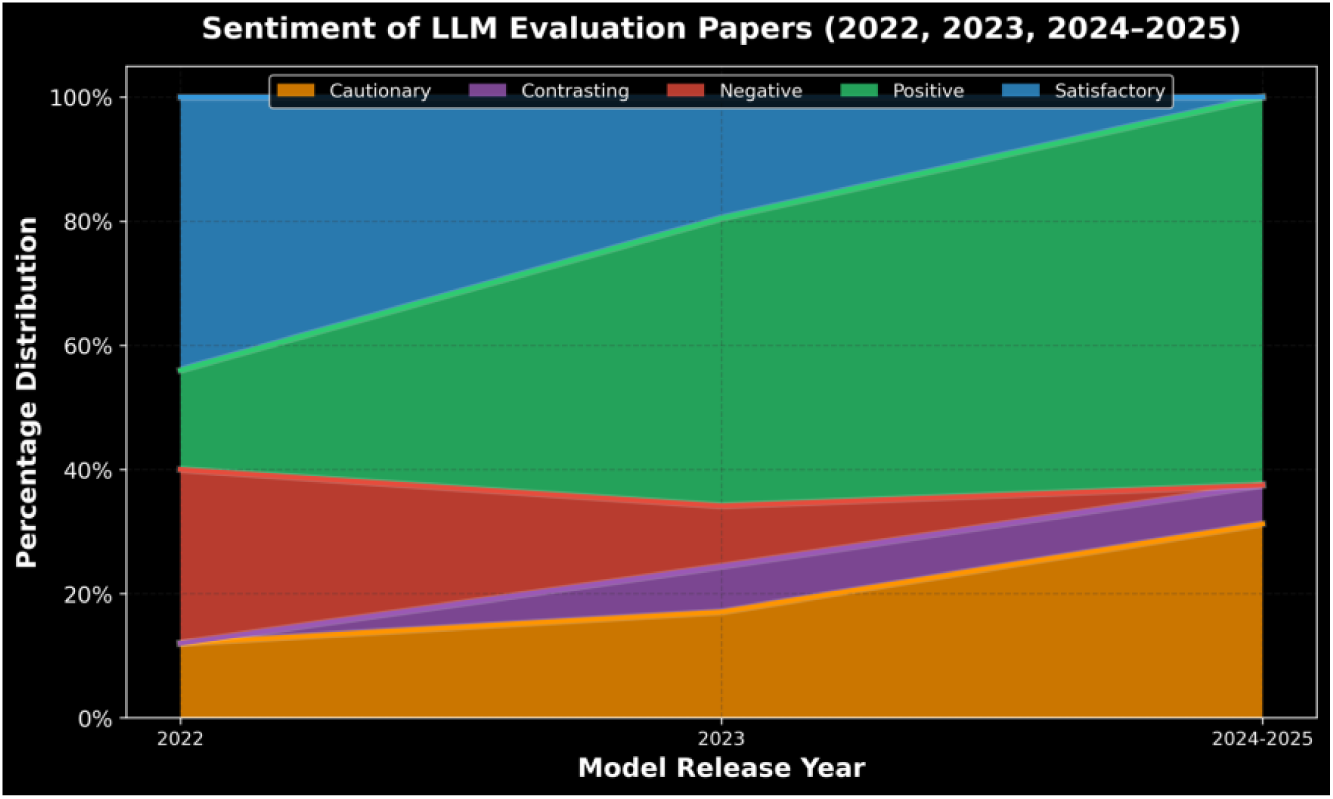
Sentiment Distribution of LLM Evaluation Papers by Model Release Year

In the pre-LLM era (2018–2021), sentiment in the literature was overwhelmingly positive. Research during this period largely centered on task-specific or fine-tuned transformer models and was typically published only after achieving favorable outcomes. The incentive to report negative or inconclusive results was minimal, and the broader evaluation landscape became dominated by confirmatory research. In practice, publishing negative findings was nearly impossible – particularly in biomedical literature, where publication bias heavily favors positive results. This dynamic changed with the emergence of general-purpose LLMs, which introduced a new norm: critiquing other models – including high-profile benchmarks – became not only acceptable but encouraged. Unlike custom, institution-specific AI systems, these models fostered a more critical research culture. A turning point came in 2022 with the release of ChatGPT 3.5, marking the rise of general-purpose LLMs. Sentiment in evaluations became more varied: approximately 40% of papers were rated Satisfactory, around 30% Negative, and fewer than 20% Positive. A new Cautionary category also appeared, highlighting concerns about safety, reliability, and alignment between model outputs and real-world expectations—especially in clinical contexts. Early evaluations often took an exploratory tone, shaped by both curiosity and a lack of methodological consensus.

By 2023, sentiment shifted again. Positive sentiment increased sharply to over 40%, while Negative sentiment declined by nearly half. Cautionary and Contrasting categories grew modestly, indicating a more nuanced reception. This optimism coincided with the release of stronger models such as GPT-4, Claude, Gemini, Mistral, and LLaMA, and likely reflects increased evaluator familiarity. However, sentiment in 2023 may have outpaced measured performance: the unweighted mean validity score was only 55%, while the weighted mean rose to 61%, driven by a handful of high-performing studies. This suggests that sentiment may have been influenced more by novelty, narrative framing, or selective task scope than by systematic empirical success. Note that sentiment was evaluated purely on language-based statements while performance was evaluated based on numerical values given in the paper. There was a very strong overall correlation (r ≈ 0.99) between average and weighted validity scores across sentiments, although “contrasting” and “cautionary” cases showed slightly more variability.

Figure 3 tracks LLM performance from 2022 through 2025, plotting both unweighted and weighted mean validity scores. A secondary dashed line shows adjusted trends that account for a 2025 study [3], which notably decreased the weighted scores due to its focus on the presence or absence of safety disclaimers, rather than diagnostic accuracy. It significantly skewed results due to the limited number of 2024–2025 evaluations. While the study raised important concerns about declining safety messaging in newer models, its methodology – based on high-throughput prompting rather than context-aware evaluation – may limit its generalizability, particularly for patient-facing use cases.

**Figure 3.**
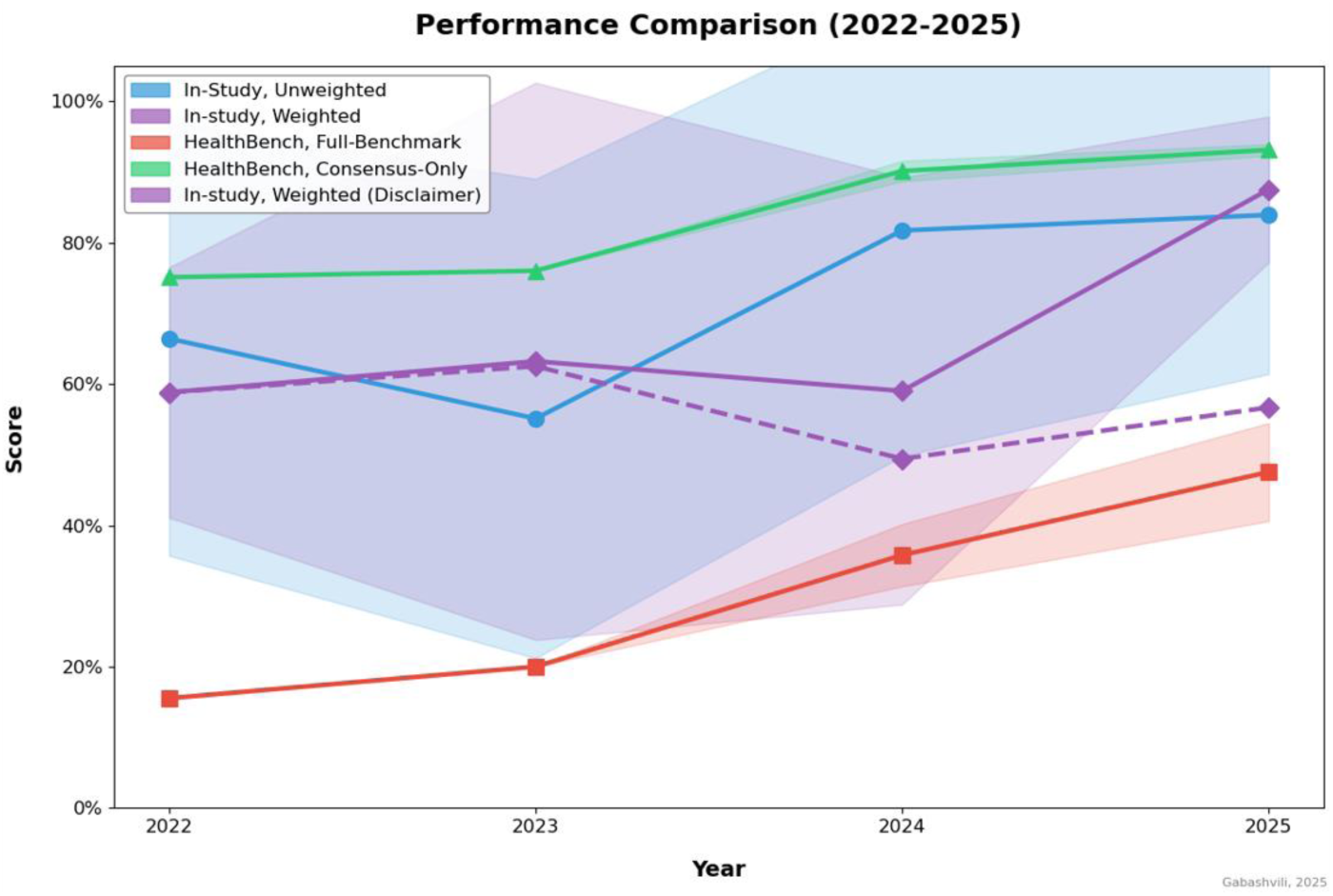
LLM Performance Trends by Model Release Year Solid lines show primary unweighted and weighted averages. The dashed purple line indicates a secondary weighted estimate including controversial data subset, as described in the text

Overall, model performance improved steadily, with both weighted and unweighted validity peaking in 2025. The unweighted mean dipped in 2023, while the weighted mean declined in 2024. This pattern likely reflects increased evaluation difficulty rather than a regression in model quality, as studies shifted toward high-stakes tasks such as image-based diagnostics, complex reasoning, multimodal interpretation, and longitudinal decision-making, with larger, more rigorous studies applying stricter scoring criteria.

One notable outlier affecting weighted scores was a 2025 ArXiv preprint [3], which assigned unusually low validity scores to newer models by evaluating medical disclaimers in LLM and VLM outputs across generations. Based on 500 dermatology image prompts, disclaimer frequency fell sharply, dropping more than an order of magnitude for both LLMs and VLMs between 2022 and 2025. Incorporating data from this paper lowers the weighted average to below 50% in 2024 and below 60% in 2025, as shown by the dashed line in Fig. 3. However, independent replication attempts failed to reproduce these results for newer releases, with evidence suggesting conditional model behavior – models appeared to suppress disclaimers once they “recognized” evaluation contexts, raising concerns about test leakage and instrumentation bias. Moreover, the study’s methodology – rapid-fire API prompts – may not accurately reflect real-world usage, especially in patient-facing environments. Effective safety assessments likely require longitudinal, context-aware simulations rather than static high-throughput testing.

This divergence emphasizes the importance of interpreting performance in light of evaluation design. Unweighted metrics often reflect early enthusiasm and exploratory testing, while weighted scores – especially those integrating low-confidence studies – offer a more cautious but arguably more representative picture. To reduce annual noise and account for overlapping publication timelines, 2024 and 2025 data were combined into a single period labeled “2024–2025” in Figure 2. This aggregated view reveals a maturing perspective: Positive sentiment remained strong (∼45%), but Cautionary papers increased, while Negative and Satisfactory studies declined. Few papers adopted a Contrasting tone, typically reserved for mixed or ambiguous findings.

A chi-square test of independence confirmed that sentiment distributions differed significantly across years (χ² = 23.46, df = 8, p = 0.0028), indicating that shifts in sentiment were statistically associated with model evolution and evaluation practices.

In sum, sentiment over time followed a U-shaped trajectory: 2022 marked initial skepticism and concern about early general-purpose LLMs. 2023 reflected a wave of optimism and enthusiasm as new models improved. 2024–2025 brought a period of reflection, critical appraisal, and methodological refinement.

This evolution reflects broader trends in AI adoption, shifting from initial hype toward more measured, task-specific integration. A recurring theme was the disconnect between AI’s outputs and evaluator expectations – especially in clinical decision-making, where subjective alignment [4], reliability, and trust are paramount. Performance steadily improved over time, with weighted mean validity reaching 88% by publication year (or 80% when including the disclaimer dataset [3]) and 87% by model release year in 2025. While no single model consistently outperformed across all domains, many exhibited specialized strengths. The ChatGPT series led in empathy, diagnostic accuracy, lesion segmentation, and anatomical localization. Gemini excelled in procedural coding, report generation, and lesion detection, while models like DeepSeek and Claude stood out in structured synthesis and reasoning. As capabilities continue to converge, performance gaps are narrowing, though disparities persist in diagnostic accuracy across skin tones, with lower performance in Fitzpatrick types III–VI compared to lighter tones. However, preliminary reevaluations with ChatGPT-5 showed measurable improvement over prior generations. Frequent model updates further complicate evaluation, rendering benchmarks quickly outdated and highlighting the ongoing challenge of assessing LLMs in a rapidly evolving landscape.

All year-to-year comparisons of weighted validity scores yielded statistically significant differences (p < 0.001), confirming that observed changes were unlikely due to random variation. Notably, scores in 2023 were significantly lower than in all other years, while 2025 scores were significantly higher, even after adjusting for sample size.

### Evaluation Methods and Benchmarking Trend

Our pooled meta-analysis, incorporating over 2,500 evaluation scores from 88 studies, revealed substantial heterogeneity in LLM performance assessments. In 2025, Cochran’s Q statistic reached 24,249.17 (df = 2,516), corresponding to an I² value of 89.6%. This indicates that nearly 90% of the observed variation is due to true differences in model behavior, evaluation design, and task difficulty, rather than sampling error.

This high level of heterogeneity reflects the diversity of LLMs, prompt formats, evaluator types, and task domains represented in our dataset. Notably, even in a more narrowly focused context – specifically, a 2025 meta-analysis of 13 studies evaluating ChatGPT’s performance on dermatology board-style exams – substantial heterogeneity was observed (I² = 72.97%). That analysis drew from over 5,000 exam-style questions, primarily composed of text-based, single-best-answer multiple-choice items [16].

While some studies focused solely on structured text prompts, others tested performance in multimodal, multilingual, or image-rich scenarios. For example, GPT-4 consistently outperformed GPT-3.5 on matched benchmarks, but the overall effect size was diluted by the increased complexity and variability of tasks introduced in later studies.

The dominant models in biomedical AI shifted markedly over time. Prior to 2023, the number of publications was small and heavily reliant on BERT variants and fine-tuned transformer models. By 2023, ChatGPT and its variants had become the most widely tested models and no specialized machine learning expertise was no longer needed for applications and evaluations. Although ChatGPT remains the most frequently evaluated model as of mid-2025, its relative dominance is declining due to the influx of newer models: our dataset includes over 50 unique LLMs spanning multiple development teams.

Evaluation metrics remained surprisingly stable across time. Accuracy persisted as the most frequently reported metric, despite the proliferation of more nuanced scoring frameworks. However, rubric-based evaluations gained ground, with increasing emphasis on dimensions such as instruction alignment, communication quality, context awareness, and clinical safety. HealthBench [5] exemplified this shift, encompassing 48,562 individual criteria rated by 262 physicians across 60 countries.

By 2024, a new paradigm emerged: recursive AI evaluation, in which AI models validated the performance of other AI systems, benchmarked against expert judgment. Nonetheless, our analysis did not detect statistically significant year-over-year increases in the use of rubrics, multimodal tasks, human-AI comparisons, or reasoning complexity. This suggests that evaluation standards were already ambitious by 2023 and have since evolved more incrementally than radically.

## Discussion

### Model Behavior, Evaluation Limitations, and Real-World Implications

This review highlights the rapid acceleration, growing complexity, and shifting evaluative paradigms in LLM research within healthcare. While model performance has measurably improved, particularly among newer reasoning-optimized systems, LLM behavior remains highly task-sensitive and context-dependent.

While newer models generally offer improved capabilities, older models can outperform them in specific contexts. For example, GPT-3.5 correctly diagnosed a ganglionic cyst that GPT-4 misclassified [6] and produced better patient education materials at a seventh-grade reading level for rare dermatologic conditions [7]. Older models are often faster, cheaper, and more consistent for straightforward tasks like summarization, data extraction, or basic content generation. They also tend to follow strict formatting or style instructions more reliably whereas newer models may deviate due to enhanced creativity, a trade-off linked to “behavioral drift,” where model responses shift over time, sometimes resulting in overthinking or less predictable behavior. These examples highlight the need to choose models based on task requirements rather than defaulting to the latest version.

Among the 50+ general-purpose models evaluated, Retrieval-Augmented Generation (RAG) abilities frequently stood out—especially in underexplored or controversial subdomains like psychodermatology. Traditional LLMs often hallucinated or failed to address conditions like PATM (“People Allergic to Me”), which lack formal medical recognition [8]. These models prioritized PubMed-indexed literature as high-quality sources, thereby overlooking substantial patient-driven research. In contrast, RAG-enabled models demonstrated better factual grounding and contextual fluency.

Perhaps most striking were the emergent behavioral capabilities exhibited by models released in the latter half of 2024, a phenomenon that persisted in 2025. Claude 3.5 Sonnet, for instance, exhibited epistemic humility – opting to express uncertainty or request clarification when unsure, rather than defaulting to hallucinated answers (Appendix 4). This represents a meaningful step forward in safety-aligned model behavior.

Specialized models like DermGPT [7] also showed strong task-specific performance, demonstrating the potential of domain-tuned architectures. While adoption of custom GPTs remains limited, personalization and context-aware fine-tuning offer promising directions. Future evaluations should explicitly account for model–user alignment, as LLMs increasingly tailor responses to individual user inputs.

Model outputs in medication management were also noteworthy. For example, in a case involving corticosteroids and anti-diabetic medications, ChatGPT-4o correctly identified hyperglycemia as the primary risk – outperforming a benchmark answer that erroneously highlighted hypoglycemia. Such examples suggest improvements not only in language understanding but also in clinical reasoning.

These performance gains have been aided by a shift in annotation practices. Industry developers increasingly rely on domain experts for labeling and evaluation, rather than crowdsourced workers. Still, benchmark datasets often remain noisy or weakly validated. For instance, widely-used datasets – such as MedQA, MedMCQA, PubMedQA, and MMLU – suffer from lack of clinical realism, insufficient validation, questionable authorship, and transparency issues [9], and up to 30% of answers in the widely cited “Humanity’s Last Exam” are flawed [10]. Our own audit revealed similar concerns in other datasets including Cognet [11] and HealthBench [5] (Appendix 4). Cross-linguistic ambiguity, regional variation, and evolving medical norms pose major challenges. In one striking example from our corpus (Appendix 3), ChatGPT-4o correctly identified corticosteroid-induced *hyperglycemia* – contradicting the benchmark’s erroneous *hypoglycemia* label – and was commended by human evaluators for its superior clinical reasoning [12]. This underscores how flawed or outdated benchmarks can obscure true model accuracy and limit their perceived relevance.

This points to a growing paradox: as models become more powerful and opaque, the transparency and trustworthiness of their evaluation pipelines may erode. The increasing use of orchestration layers, synthetic inputs, and modular reasoning paths makes it harder to audit how conclusions are generated – posing new challenges for reproducibility, regulation, and informed adoption.

A crucial finding from 2025 centers on the gap between model competence and real-world usability. Bean et al. [13] found that while GPT-4o, LLaMA 3, and Command R+ achieved ∼95% benchmark accuracy in diagnostic tasks, actual users interacting with these models performed no better than control groups correctly identifying conditions only 34.5% of the time and selecting appropriate next steps just 44.2% of the time. This highlights a fundamental limitation of current LLMs: benchmark performance does not guarantee real-world utility.

Human-AI misalignment, especially in emotionally charged or ambiguous queries, could drive suboptimal outcomes. The inability of LLMs to detect when they fail to understand a user’s true needs and true intent is especially problematic in healthcare settings, where next-step decisions are critical. Bean et al.’s study is a timely reminder: if LLMs are built for human interaction, they must be evaluated with humans, not merely on humans.

To date, no published research has fully contradicted those findings for unassisted layperson usage in real world.

Looking forward, the LLM ecosystem is trending toward more modular, hybrid architectures. These include RAG-based querying for factual precision, vision modules for image– and video-based diagnostics, and agentic frameworks for reasoning over multiple steps.

Such systems may improve factuality and adaptability while reducing hallucination risks. However, success will hinge on transparent testing, robust human-in-the-loop design, and ongoing monitoring of user-centered outcomes.

Recent studies reflect both the promise and the peril of these tools. Brodeur et al. [14] found that OpenAI’s o1-preview model exhibited superhuman reasoning in diagnosis and management planning, motivating the urgent need for prospective trials. When paired with OpenAI’s o3 model, a multi-agent orchestration framework emulating a collaborative panel of virtual doctors, solved clinical diagnosis tasks more effectively than off-the-shelf LLMs or average individual generalist physicians (4 times higher diagnostic accuracy [15]).

Together, these findings underscore a critical tension: LLMs are evolving quickly, but their clinical integration must be measured, transparent, and grounded in rigorous, user-aware testing frameworks.

### Implications for Future Evaluation Frameworks

As general-purpose LLMs become increasingly integrated into health-related workflows, the evaluation landscape must evolve in tandem – not only to capture raw performance, but to assess models’ safety, alignment, and usability in complex, high-stakes environments. This review highlights several structural shifts already underway, and outlines essential directions for the next generation of LLM evaluation frameworks.

Many current evaluations still rely on static benchmarks and single-turn prompt–response formats. While useful for early comparisons, such methods increasingly fail to reflect how LLMs are used in practice—particularly in clinical and emotionally complex scenarios where users seek clarification, ask follow-ups, or shift intent mid-conversation. As Bean et al. [13] demonstrated, benchmark success does not translate directly to successful real-world outcomes, especially when lay users are involved.

Future evaluations must therefore move toward interactive, user-in-the-loop testing, where model performance is assessed not just on isolated answers, but on its ability to engage productively over time. This requires simulation environments, standardized user scenarios, and metrics that reward dialog coherence, intent recognition, and adaptive behavior—not just factual recall.

The rise of contrasting sentiment in 2025 reflects growing awareness that evaluator effects (such as human vs. AI graders, prompt phrasing, or rubric sensitivity) can substantially influence reported outcomes. Papers using the same model often reach divergent conclusions based on who evaluates, how, and on what criteria.

To counteract this, future frameworks must adopt evaluator-aware designs with transparent documentation of grading processes, inclusion of both domain experts and lay evaluators, consensus scoring and meta-evaluation of the evaluators themselves. Without this, performance assessments risk being as subjective and brittle as the models they aim to judge.

As LLMs become increasingly personalized, offering different responses based on user history, expertise level, or regional context, evaluations must adapt. A correct answer for a medical student may differ from the correct level of explanation for a patient. Similarly, safety-critical recommendations may need to vary based on jurisdiction or availability of care.

Evaluation frameworks must begin treating context sensitivity and personalization not as confounds, but as necessary features to assess. Scoring should capture whether models adjust tone, uncertainty, or actionability based on user profile or expressed needs—while still meeting expert-derived safety and accuracy thresholds.

Emerging architectures are no longer monolithic text generators but modular systems combining Retrieval-Augmented Generation (RAG) for factual grounding, vision modules, symbolic tools for calculation and data manipulation, and multi-agent orchestration layers for sequential planning.

Future evaluation frameworks must assess whole-system behavior and not just module-level correctness. This will likely require scenario-based simulations, synthetic test environments, and real-world deployment auditing.

Summarizing, the field of LLM evaluation is entering a new phase—defined less by model capabilities and more by methodological rigor, human-centered validation, and systemic accountability. The next generation of evaluation frameworks must reflect the way people actually use LLMs, account for personalization, context, and collaboration, adapt to hybrid, multi-agent, and tool-augmented architectures, and prioritize data quality, evaluator transparency, and reproducibility.

Only by embracing these challenges can we ensure that LLMs are not just accurate but aligned with real-world needs in medicine and beyond.

## Conclusion

As LLMs become integrated into healthcare workflows, their growing capabilities demand a parallel evolution in how we evaluate, monitor, and trust them. Our systematic review – spanning over 2,500 evaluation scores from 88 studies – confirms steady improvements in reasoning, factual grounding, and context sensitivity. However, these gains remain uneven, task-specific, and often compromised by persistent challenges such as hallucinations, evaluator bias, and disconnects between benchmark performance and real-world utility.

The rapid turnover of state-of-the-art models has drastically shortened the relevance window for evaluations. By mid-2025, two-thirds of the model assessments in this review concerned already-deprecated systems. The launch of ChatGPT 5 on August 7 2025 pushed that share to 87%.

This pace highlights a widening gap between the cycles of model development and traditional academic publication, underscoring the need for dynamic, living evaluation strategies and real-time benchmarking infrastructure.

Sentiment and performance patterns from 2022 to 2025 reveal a dynamic evaluative landscape – transitioning from initial caution, through a peak of enthusiasm, into a more critical and nuanced phase. In particular, the rise of “Contrasting” sentiment in 2025 signals a maturing recognition that accuracy alone is insufficient. Evaluations must consider *who* is assessing the model, *how* they are doing so, and *under what conditions*. These shifts call for evaluator-aware frameworks that treat human feedback not as noise, but as a meaningful and contextual signal.

Emerging model behaviors – such as epistemic humility, adaptive clarification, and personalized tone – mark an inflection point in LLM-human interaction. Hybrid approaches, including Retrieval-Augmented Generation (RAG) and modular architectures, have shown promise in high-complexity tasks like rare disease detection and medication reconciliation. Yet even highly capable models can fail in practice when not designed with human-centered principles.

To truly support patients, clinicians, and healthcare systems, future evaluations must move beyond static benchmarks. They should simulate real-world use: patient scenarios, longitudinal interactions, multi-agent collaboration, and clinical decision-making under uncertainty. Evaluation frameworks must prioritize not only technical performance but also usability, interpretability, and adaptability to clinical contexts.

As LLMs move closer to real-world patient care, their design and evaluation must reflect the complexity of the people and systems they aim to support. These tools should not only answer questions – they should earn trust, adapt to context, and respect the stakes involved.

## Supporting information

Appendix 1: Database Search Strategies and Query Specifications

Appendix 2: Final Selection of Papers from 2023 review annotated a year later

Appendix 3: Final Selection of Papers from this systematic review

Appendix 4: Evaluation Scores

## Acknowledgements

The author acknowledges the assistance of ChatGPT, Gemini, Claude, and other models for their contributions to code base, stylistic refinement, and grammar enhancement.

## Data Availability Statement

Lists of all papers selected for final analysis in 2023 and 2025, along with over 2,500 extracted scores, are provided in the appendices, which serve as the supplementary materials.

## Funding

This research received no external funding.

## Conflicts of Interest

None declared.

## Appendices

**Appendix 1**. Database Search Strategies.

Live, regularly updated editions are available at OSF: https://osf.io/7jxek

**Appendix 2**. Annotated Papers from 2023.

Condensed table provided in the Supplementary Materials; full dataset available at OSF: https://osf.io/98rmw)

**Appendix 3**. Annotated Papers from 2025. Condensed table provided in the Supplementary Materials; full dataset available at OSF: https://osf.io/fsa4q)

**Appendix 4**. Evaluation Scores. Complete dataset available at OSF: https://osf.io/gbwam)

## Notes

### Competing Interest Statement

The authors have declared no competing interest.

### Clinical Protocols

https://www.crd.york.ac.uk/PROSPERO/view/CRD42023417336

### Funding Statement

This study did not receive any funding

## References

1. Gabashvili IS. ChatGPT in dermatology: a comprehensive systematic review. medRxiv. 2023 Jun 12:2023–06. 10.1101/2023.06.11.23291252

2. Gabashvili, IS. The Impact and Applications of General-Purpose AI Tools Across Industries and Disciplines. OSF. Created May 5, 2023; last updated August 10, 2025. doi:10.17605/OSF.IO/87U6Q.

3. Sharma S, Alaa AM, Daneshjou R. A Systematic Analysis of Declining Medical Safety Messaging in Generative AI Models. arXiv preprint arXiv:2507.08030 [cs.CL]. 2025 Jul 8. 10.48550/arXiv.2507.08030

4. Sengupta D, Panda S. Divergent Realities: A Comparative Analysis of Human Expert vs. Artificial Intelligence Based Generation and Evaluation of Treatment Plans in Dermatology. arXiv preprint arXiv:2507.05716. 2025 Jul 8. 10.48550/arXiv.2507.05716

5. Arora RK, Wei J, Hicks RS, Bowman P, Quiñonero-Candela J, Tsimpourlas F, Sharman M, Shah M, Vallone A, Beutel A, Heidecke J. Healthbench: Evaluating large language models towards improved human health. arXiv preprint arXiv:2505.08775. 2025 May 13. 10.48550/arXiv.2505.08775

6. Manoharan P, Surapaneni KM. Assessing the diagnostic capability of ChatGPT through clinical case scenarios in dermatology. Indian J Dermatol Venereol Leprol. 2024 May 25:1–3. doi: 10.25259/IJDVL_1267_2023. Epub ahead of print. PMID: 38841923.

7. Lambert R, Choo ZY, Gradwohl K, Schroedl L, Ruiz De Luzuriaga A. Assessing the Application of Large Language Models in Generating Dermatologic Patient Education Materials According to Reading Level: Qualitative Study. JMIR Dermatol. 2024 May 16;7:e55898. doi: 10.2196/55898. PMID: 38754096; PMCID: PMC11140271.

8. Gabashvili IS. Cutaneous Bacteria in the Gut Microbiome as Biomarkers of Systemic Malodor and People Are Allergic to Me (PATM) Conditions: Insights From a Virtually Conducted Clinical Trial. JMIR Dermatol 2020;3(1):e10508 doi: 10.2196/10508

9. Alwakeel M, Nagori A, Krishnamoorthy V, Kamaleswaran R. Evaluating LLMs in Medicine: A Call for Rigor, Transparency. arXiv preprint arXiv:2507.08916. 2025 Jul 11. 10.48550/arXiv.2507.08916

10. Skarlinski, Michael, Jon Laurent, Albert Bou, and Andrew White. About 30% of Humanity’s Last Exam Chemistry/Biology Answers Are Likely Wrong. White paper. Future House Institute, July 23, 2025. https://www.futurehouse.org/research-announcements/hle-exam.

11. Panagoulias, D.P. et al. (2025). COGnitive Network Evaluation Toolkit for Medical Domains: Evaluating Large Language Model Performance in Medical Diagnostics—An Assessment Framework and Dataset for Healthcare AI. In: Nagar, A., Jat, D.S., Mishra, D., Joshi, A. (eds) Intelligent Sustainable Systems. Worlds4 2024. Lecture Notes in Networks and Systems, vol 1178. Springer, Singapore. 10.1007/978-981-97-9559-8_38 https://huggingface.co/datasets/DimitriosPanagoulias/COGNET-MD/viewer/

12. Jonathan Shapiro, Sharon Baum, Felix Pavlotzky, Yaron Ben Mordehai, Aviv Barzilai, Tamar Freud, Rotem Gershon, Application of a natural language processing artificial intelligence tool in psoriasis: A cross-sectional comparative study on identifying affected areas in patients’ data, Clinics in Dermatology, Volume 42, Issue 5, 2024, Pages 480–486, ISSN 0738-081X, 10.1016/j.clindermatol.2024.06.018. https://www.sciencedirect.com/science/article/pii/S0738081X24001020

13. Bean AM, Payne R, Parsons G, Kirk HR, Ciro J, Mosquera R, Monsalve SH, Ekanayaka AS, Tarassenko L, Rocher L, Mahdi A. Clinical knowledge in LLMs does not translate to human interactions. arXiv preprint arXiv:2504.18919. 2025 Apr 26. 10.48550/arXiv.2504.18919

14. Brodeur PG, Buckley TA, Kanjee Z, Goh E, Ling EB, Jain P, Cabral S, Abdulnour RE, Haimovich AD, Freed JA, Olson A. Superhuman performance of a large language model on the reasoning tasks of a physician. arXiv preprint arXiv:2412.10849. 2024 Dec 14. Latest version: Mon, 2 Jun 2025 20:29:39 UTC

15. Nori H, Daswani M, Kelly C, Lundberg S, Ribeiro MT, Wilson M, Liu X, Sounderajah V, Carlson J, Lungren MP, Gross B. Sequential Diagnosis with Language Models. arXiv preprint arXiv:2506.22405. 2025 Jun 27. Latest version: Wed, 2 Jul 2025 17:58:37 UTC

16. Andrew, A. A meta-analysis of ChatGPT’s performance on dermatology specialty-level (board-style) certification questions. Indian Dermatology Online Journal. 2025, July 23. 10.4103/idoj.idoj_1250_24

