## Appendix 1: Database Search Strategies and Query Specifications for "Evaluating General-Purpose LLMs for Patient-Facing Use: Dermatology-Centered Systematic Review and Meta-Analysis"

### Appendix 1. Search Strategy

Repositories searched: EuropePMC, Semantic Scholar, PubMed, Dimensions AI, MedRxiv, BioRxiv, Google Scholar, arXiv, and Research Square.

For arXiv and Semantic Scholar, both the web interface and API were used (implementation code: https://osf.io/gekuf, https://osf.io/hd2at). Since arXiv’s native search is limited to titles, abstracts, authors, and metadata (not full text), combining the arXiv API with Dimensions.AI yielded a more comprehensive set of arXiv-hosted papers. All matches were manually reviewed for relevance to dermatology/AI/ML context.

| Database / Repository | Search Query (Simplified) | Filters / Limits | Notes |
| --- | --- | --- | --- |
| PubMed | (Dermatology OR skin OR hair OR nails OR dermatitis OR acne OR vitiligo OR alopecia OR "lichen planus" OR cosmetics OR eczema OR malodor OR hyperhidrosis OR bromhidrosis OR "olfactory reference syndrome" OR rosacea OR sunburn) AND ("ChatGPT" OR "chat GPT") | Additional queries restricted to specific journal ISSNs | Used MeSH terms where available |
| Semantic Scholar | (Dermatology OR skin OR hair OR nails OR dermatitis OR acne OR vitiligo OR alopecia OR "lichen planus" OR cosmetics OR eczema OR "body odor" OR "excessive sweating" OR "olfactory reference syndrome" OR rosacea OR sunburn) AND ChatGPT | Publication years: 2022–2025 | Different handling between web UI and API |
| MedRxiv / BioRxiv | dermatology AND chatgpt | No year filter | Direct search via site interface |
| EuropePMC | (dermatology OR skin OR hair OR nails OR dermatitis OR acne OR vitiligo OR alopecia OR "lichen planus" OR cosmetics OR eczema OR malodor OR hyperhidrosis OR bromhidrosis OR "olfactory reference syndrome" OR rosacea OR sunburn) AND (ChatGPT OR "chat GPT") | Keyword (KW) search yields fewer results | Tested both full-text and keyword searches |
| arXiv | Dermatology terms list + ("ChatGPT" OR "chat GPT") | No year filter | Used API and generated URL-encoded query strings |
| Google Scholar | ("Dermatology" OR "skin conditions" OR "hair disorders" OR "nail disorders" OR "dermatitis" OR "acne" OR "vitiligo" OR "alopecia" OR "lichen planus" OR "cosmetics" OR "eczema" OR "malodor" OR "hyperhidrosis" OR "bromhidrosis" OR "olfactory reference syndrome" OR "rosacea" OR "sunburn") AND ("ChatGPT" OR "chat GPT") | Publication years: 2022–2025 | Standard interface search |
| Dimensions.AI | (dermatology OR skin OR hair OR nails OR dermatitis OR acne OR vitiligo OR alopecia OR "lichen planus" OR cosmetics OR eczema OR malodor OR hyperhidrosis OR bromhidrosis OR "olfactory reference syndrome" OR rosacea OR sunburn) AND ("ChatGPT" OR "chat GPT") | Publication years: 2022–2025 | Final 2025 search was expanded to include more LLMs and restricted to title and abstracts: (Dermatologic OR Dermatology) AND ("language model" OR ChatGPT OR (Chat GPT) OR ChatGPT4 OR transformer) |
| Research Square | (dermatology OR skin OR hair OR nails OR dermatitis OR acne OR vitiligo OR alopecia OR "lichen planus" OR cosmetics OR eczema OR malodor OR hyperhidrosis OR bromhidrosis OR "olfactory reference syndrome" OR rosacea OR sunburn) AND (ChatGPT OR "chat GPT") | Publication years: 2022–2025 | [Example link](https://www.researchsquare.com/browse?offset=0&postedAfter=2022-01-01&status=all&title=dermatology%20AND%20chatgpt) for title and abstract search  [Link](https://www.researchsquare.com/browse?offset=0&postedAfter=2022-01-01&status=all&unified=dermatology%20AND%20chatgpt) for full search  Final 2025 search was expanded to include more LLMs and restricted to title and abstracts. |
| General AI Search for latest models (2025) | "Deepseek R1" OR "ChatGPT o3" OR "Gemini 2.5" OR "ChatGPT o4" OR "ChatGPT 4.1" OR “ChatGPT 5” OR "Claude 4" OR "Grok 3" OR "Grok 4" | Publication year: 2025; Fields of Research: 32, 42 | Applied to Dimensions.AI |
